# Vaccine effectiveness of the BNT162b2 mRNA COVID-19 vaccine against RT-PCR confirmed SARS-CoV-2 infections, hospitalisations and mortality in prioritised risk groups

**DOI:** 10.1101/2021.05.27.21257583

**Authors:** Hanne-Dorthe Emborg, Palle Valentiner-Branth, Astrid Blicher Schelde, Katrine Finderup Nielsen, Mie Agermose Gram, Ida Rask Moustsen-Helms, Manon Chaine, Ulla Holten Seidelin, Jens Nielsen

**Author notes:** Corresponding Author Hanne-Dorthe Emborg, Department of Infectious Disease Epidemiology and Prevention, Statens Serum Institut, 5 Artillerivej 2300 Copenhagen, Denmark.

## Abstract

**Objective:** To estimate in a real life setting, the vaccine effectiveness of the BNT162b2 mRNA vaccine against confirmed SARS-CoV-2 infection, hospital admission, and death among five priority groups for vaccination

**Design:** Cohort study

**Setting:** Roll-out of the BNT162b2 mRNA vaccine in Denmark

**Participants:** 864,096 individuals who were first inline to receive the BNT162b2 mRNA vaccine: 46,101 long-term care facility (LTCF) residents, 61,805 individuals 65 years and older living at home but requiring practical help and personal care (65PHC), 98,533 individuals ≥85 years of age (+85), 425,799 health-care workers (HCWs), and 231,858 individuals with comorbidities that predispose for severe COVID-19 disease (SCD).

**Intervention:** vaccination with BNT162b2 mRNA vaccine

**Main outcome measures:** RT-PCR confirmed SARS-CoV-2 infections, COVID-19 related admissions within 14 days after a confirmed SARS-CoV-2 infection, all-cause admission, COVID-19 related death within 30 days after confirmed SARS-CoV-2 infection, and all-cause death.

**Results:** Beyond 7 days after the second dose, the VE against SARS-CoV-2 infection in all groups ranged from 53-86%. In 65PHC, HCW and SCD, we observed a substantial reduction in risk of infection 0-7 days after the second dose ranging from 46-71%. The VE against COVID-19 related admissions ranged from 75-87% in all groups except +85 and HCWs where no events occurred. For COVID-19 related deaths, a significant VE was observed in LTCF residents (VE of 89%) and 65PHC (VE of 97%), whereas no events were observed in the three remaining groups. VE against all-cause death ranged from 26-73% in all groups except HCW where an insignificant VE was estimated. For all-cause admission, the VE ranged from 37-50% in all groups except in SCD where a negative VE was observed.

**Conclusion:** In a real-life setting and more than 7 days after the second dose of BNT162b2 mRNA was administered to the most vulnerable individuals, the vaccine was associated with a reduction of SARS-CoV-2 infection (53-86%) and COVID-19 related admissions (≥75%) or deaths (≥89%).

## Introduction

The efficacy of COVID-19 vaccines has been investigated in randomized controlled trials. However, the individuals included in these trials do not necessarily reflect those who are in most need of vaccination against COVID-19 in a real-life setting [1]. The Danish population was divided into vaccination priority groups that were offered COVID-19 vaccination based on various factors that could influence the risk of being infected with SARS-CoV-2 and developing severe COVID-19 disease. The COVID-19 vaccination programme was initiated on December 27, 2020, when the Pfizer-BioNTech mRNA vaccine, BNT162b2, became available. The first priority groups for vaccinations were residents in long-term care facilities (LTCF) and frontline healthcare workers (HCW). Seven days after the second dose the preliminary vaccine effectiveness (VE) estimates in preventing SARS-CoV-2 infections were 64% (95% confidence interval (CI); 14-84) and 90% (95% CI; 82-95) in these two groups, respectively [2].

The priority groups next in line for COVID-19 vaccination were i) individuals 65 years and older living at home but requiring practical help and personal care (65PHC), ii) individuals 85 years and older (+85), and iii) individuals with comorbidities that predispose for severe COVID-19 disease (SCD). From the January 6, 2021 the enrolment of 65PHC and SCD was initiated, while +85 were enrolled from February 10, 2021 (Figure 1). Real-world data on COVID-19 VE against SARS-CoV-2 infections are important; however, from a public health perspective it is also important to quantify how effective the vaccines are in preventing hospital admissions and death, in particular among the most vulnerable groups.

**Figure 1.**
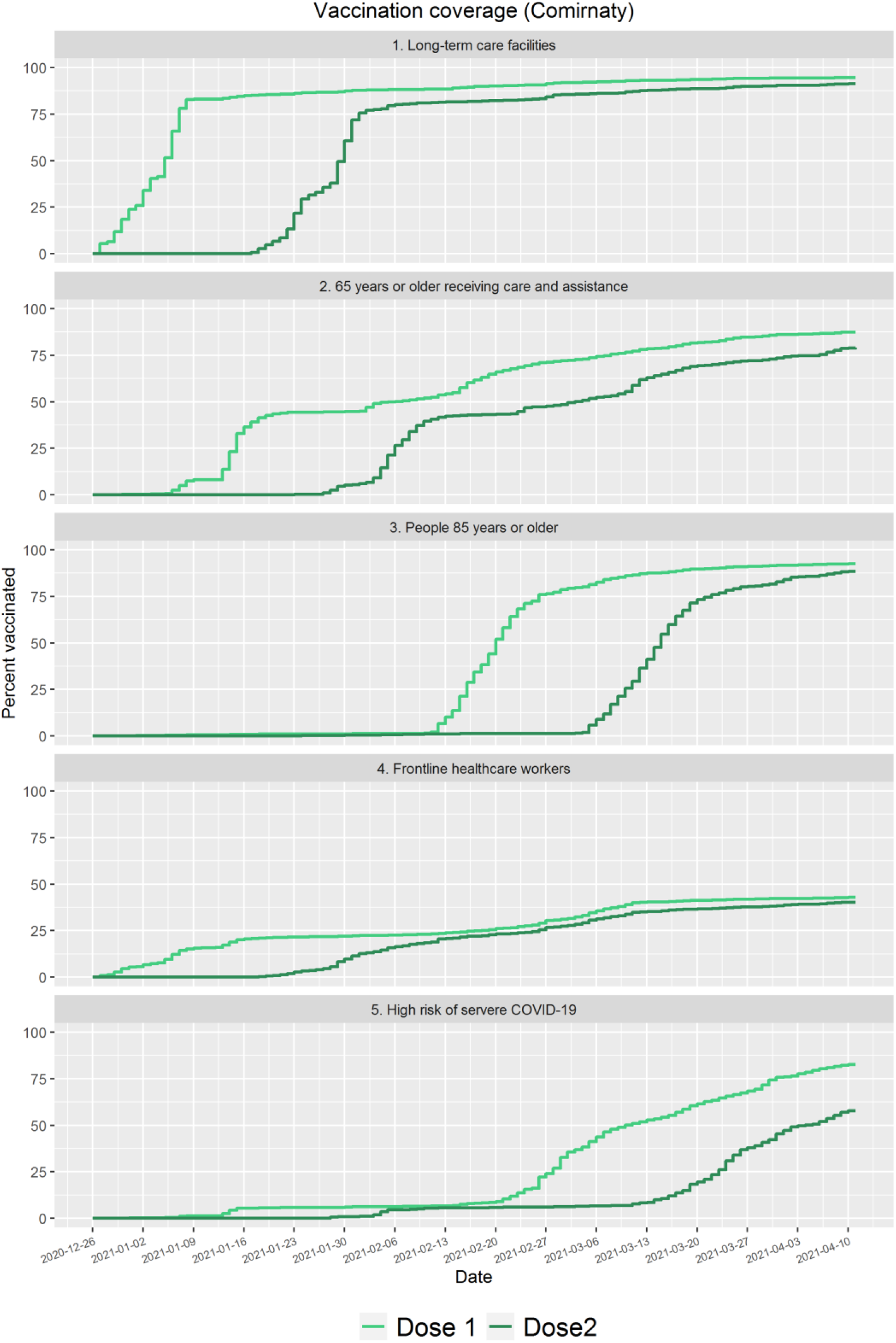
Cumulative BNT162b2 mRNA vaccine coverage for the first and second dose during the study period for each priority group

The aim of this study was to estimate BNT162b2 mRNA VE against infection with SARS-CoV-2, all-cause and COVID-19 related admissions to hospitals and death in the five priority groups LTCF residents, 65PHC, HCW, SCD, and +85.

## Methods

### Data sources

All individuals living in Denmark are registered in The Danish Civil Registration System with a unique identifier allowing individual-level linkage between various registries [3]. From this registry, we retrieved information on date of birth, immigration and emigration status, sex, and vital status. Laboratory confirmed SARS-CoV-2 infections identified with real-time Reverse Transcription Polymerase Chain Reaction (RT-PCR) are registered in the National Danish Microbiology Database (MiBa) including date of sample [4]. Information on all administered COVID-19 vaccines was retrieved from the Danish Vaccination Registry including the date the vaccine was administered, brand, and dose [5]. All hospital admissions are registered in the Danish National Patient Registry with admission and discharge dates and diagnoses coded according to the International Classification of Diseases, 10th revision (ICD-10) [6]. Comorbidity (yes/no) in the previous five years (from 2016 to 2020) was defined by diagnose codes registered for all hospital admissions. LTCF residents were identified through two sources; by linking addresses in the Danish Civil Registration System to addresses for LTCF from the Danish Health and Data Authority and through lists with unique identifiers for current LTCF residents provided by the municipalities [7,8]. Information on persons working in the healthcare sector was retrieved from The Danish Agency for Labour Market and Recruitment [9,10]. The municipalities identified individuals belonging to the priority group 65PHC and clinicians identified SCD individuals.

### Analyses

Five cohorts were defined for each of the five priority groups. Individuals in the cohorts were either living in Denmark at start of the vaccination programme (December 27, 2020) or immigrated before the end of study on April 11, 2021. For each cohort, we estimated VE relative to unvaccinated individuals on five types of events: i) confirmed SARS-CoV-2 infection, ii) all-cause admission, iii) COVID-19 related admission to hospital defined as an admission within 14 days after a confirmed SARS-CoV-2 infection, iv) all-cause death, and v) COVID-19 related death defined as death within 30 days after confirmed SARS-CoV-2 infection. For all events, the cohorts were followed from the beginning of the vaccination programme, December 27, 2020, and until receiving a dose of mRNA-1273 (Moderna) or ChAdOx1 (AstraZeneca) vaccine, emigration, death or end of follow-up (April 11, 2021), whichever came first.

Hospital admission was considered a recurrent event and defined as at least 12 hours in a hospital. Coherent admissions by date were considered as one admission. Individuals with an RT-PCR confirmed SARS-CoV-2 infection before December 27, 2020 were excluded.

The summary findings for BNT162b2 mRNA presented by the European Medicines Agency were used to identify relevant time periods of exposure after vaccination [1,11]: 1) 0-14 days after the first dose (no protection). 2) >14 days after the first dose and until the second dose (partial protection). 3) 0-7 days after the second dose (partial protection), and 4) >7 days after the second dose (highest protection). The BNT162b2 mRNA vaccine was available from December 27, 2020, the mRNA-1273 (Moderna) vaccine from January 14, 2021 and ChAdOx1 (AstraZeneca) from February 9, 2021.

For each event, the four time periods were included as time-varying exposures in a Cox regression estimating the hazard rates in vaccinated relative to unvaccinated individuals. VE was calculated as one minus the hazard ratio. The infection pressure varied considerably through the study period [2], thus calendar time was used as the underlying time in the Cox regression model. Age, sex and comorbidity were included as confounders. For confirmed SARS-CoV-2 infection, all-cause death and COVID-19 related death, admitted to hospital was also a confounder.

COVID-19 related admissions or deaths were defined as occurring within 14 or 30 days after a positive SARS-CoV-2 test, respectively. The vaccination status of an individual may change within this time, e.g. from having a positive test while unvaccinated and dying after having received the first dose, but within 30 days after the positive test. We considered this as a change in risk and included vaccination status at the time of a SARS-CoV-2 positive test as a covariate.

Data was analysed using R version 4.0.3 (R Foundation for Statistical Computing, https://www.R-project.org/).

## Results

### Descriptive data

In total 864,096 individuals were included in the priority groups LTCF residents (46,101), 65PHC (61,805), +85 (98,533), HCW (425.799), and SCD individuals (231.335) (Table 1). In all priority groups, except SCD, females were more prevalent and HCW were the youngest group, followed by SCD (Table 1).

**Table 1.**
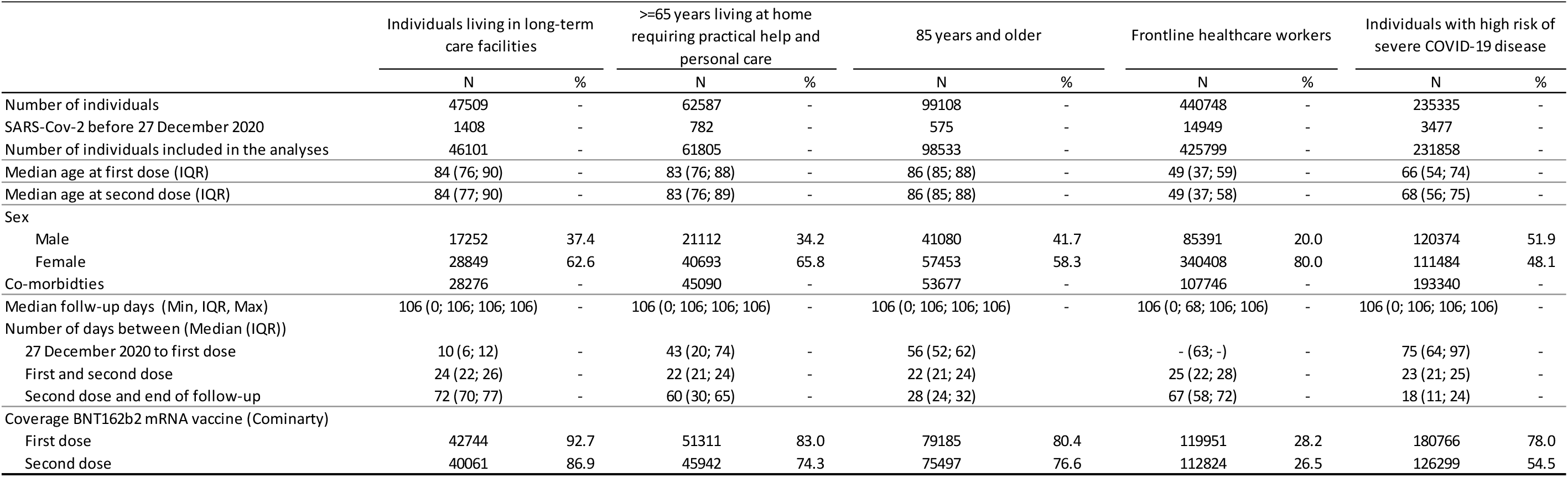
Numbers and proportions of individuals vaccinated with first and second dose BNT162b2 in five priority groups in Denmark (Dec 27 to April 11, 2021)

|  | Individuals living in long-term care facilities |  | >=65 years living at home requiring practical help and personal care |  | 85 years and older |  | Frontline healthcare workers |  | Individuals with high risk of severe COVID-19 disease |  |
| --- | --- | --- | --- | --- | --- | --- | --- | --- | --- | --- |
|  | N | % | N | % | N | % | N | % | N | % |
| Number of individuals | 47509 | - | 62587 | - | 99108 | - | 440748 | - | 235335 | - |
| SARS-Cov-2 before 27 December 2020 | 1408 | - | 782 | - | 575 | - | 14949 | - | 3477 | - |
| Number of individuals included in the analyses | 46101 | - | 61805 | - | 98533 | - | 425799 | - | 231858 | - |
| Median age at first dose (IQR) | 84 (76; 90) | - | 83 (76; 88) | - | 86 (85; 88) | - | 49 (37; 59) | - | 66 (54; 74) | - |
| Median age at second dose (IQR) | 84 (77; 90) | - | 83 (76; 89) | - | 86 (85; 88) | - | 49 (37; 58) | - | 68 (56; 75) | - |
| Sex |  |  |  |  |  |  |  |  |  |  |
| Male | 17252 | 37.4 | 21112 | 34.2 | 41080 | 41.7 | 85391 | 20.0 | 120374 | 51.9 |
| Female | 28849 | 62.6 | 40693 | 65.8 | 57453 | 58.3 | 340408 | 80.0 | 111484 | 48.1 |
| Co-morbidities | 28276 | - | 45090 | - | 53677 | - | 107746 | - | 193340 | - |
| Median follow-up days (Min, IQR, Max) | 106 (0; 106; 106; 106) | - | 106 (0; 106; 106; 106) | - | 106 (0; 106; 106; 106) | - | 106 (0; 68; 106; 106) | - | 106 (0; 106; 106; 106) | - |
| Number of days between (Median (IQR)) |  |  |  |  |  |  |  |  |  |  |
| 27 December 2020 to first dose | 10 (6; 12) | - | 43 (20; 74) | - | 56 (52; 62) | - | - (63; -) | - | 75 (64; 97) | - |
| First and second dose | 24 (22; 26) | - | 22 (21; 24) | - | 22 (21; 24) | - | 25 (22; 28) | - | 23 (21; 25) | - |
| Second dose and end of follow-up | 72 (70; 77) | - | 60 (30; 65) | - | 28 (24; 32) | - | 67 (58; 72) | - | 18 (11; 24) | - |
| Coverage BNT162b2 mRNA vaccine (Cominarty) |  |  |  |  |  |  |  |  |  |  |
| First dose | 42744 | 92.7 | 51311 | 83.0 | 79185 | 80.4 | 119951 | 28.2 | 180766 | 78.0 |
| Second dose | 40061 | 86.9 | 45942 | 74.3 | 75497 | 76.6 | 112824 | 26.5 | 126299 | 54.5 |

BNT162b2 mRNA vaccine coverage per dose by the end of the study (April 11, 2021) within each priority group is presented in Table 1, while the cumulative coverage per dose through the study period is shown in Figure 1. In LTCF residents and +85, >75% coverage was reached in both groups within 2 weeks after the vaccines were rolled out. Similarly, a high coverage for the second dose was reached within 2-3 weeks. In the remaining three priority groups, a more gradual increase in coverage was observed.

### VE against SARS-CoV-2 infection

In all five priority groups, the adjusted VE estimates were lower than the unadjusted VE estimates (Table 2). In particular, calendar time had a large impact on the adjusted estimates.

**Table 2.**
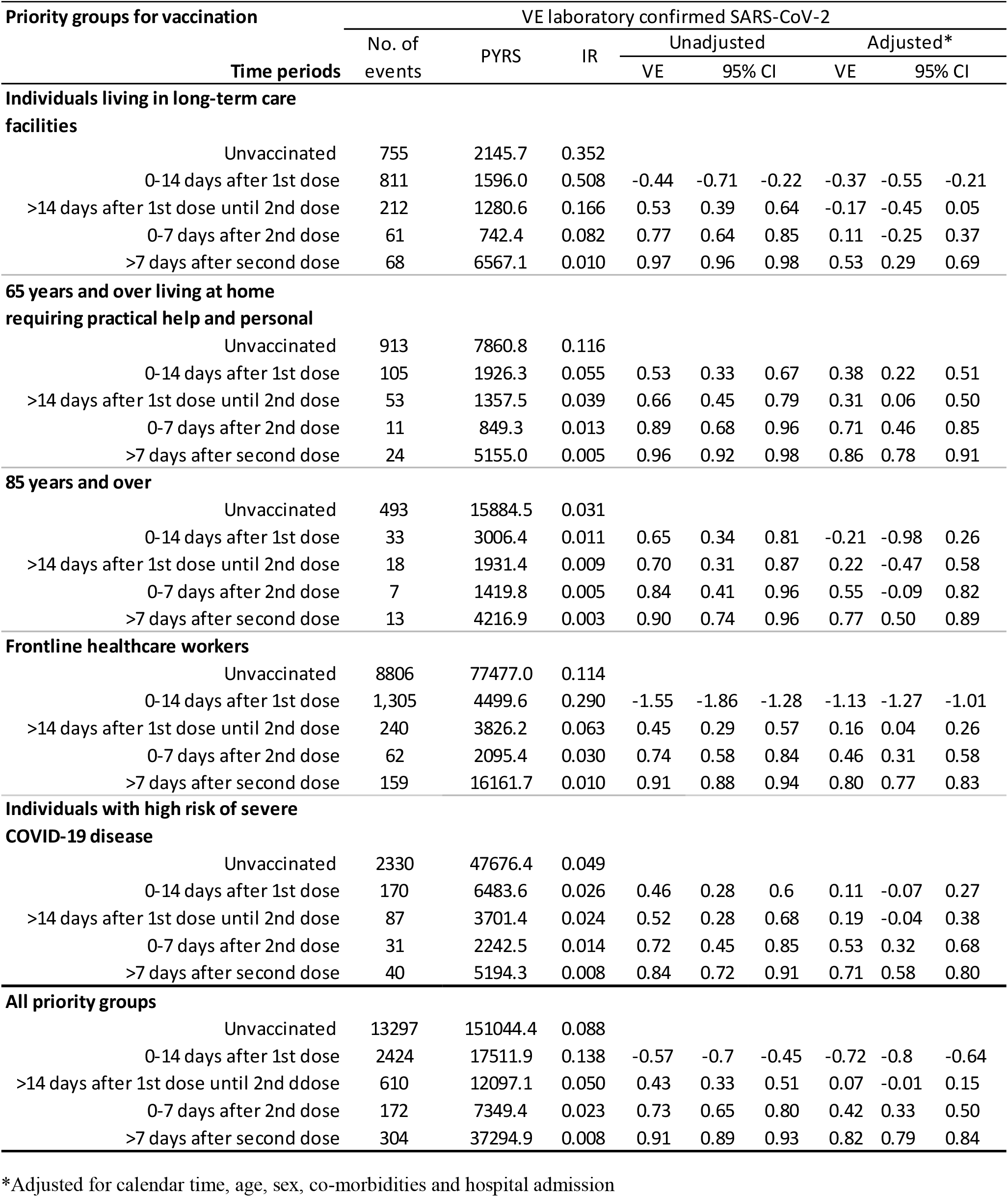
BNT162b2 mRNA vaccine effectiveness in the priority groups long term care facility residents, individuals 65 years and above requiring practical help and personal care, individuals 85 years and older, frontline healthcare workers and individuals with high risk of severe COVID-19 disease

Significant VE estimates were observed >7 days after the second BNT162b2 mRNA dose. In all five priority groups with 53% (95% CI: 29; 69) in LTCF residents, 86% (95% CI: 78; 91) in 65PHC, 77% (95% CI: 50; 89) in +85, 80% (95% CI: 77; 83) in HCW and 71% (95% CI: 58; 80) in SCD. In addition, significant protective VE estimates were observed from 14 days until the second dose in HCW of 16% (95% CI: 4; 26), from 0-7 days after the second dose in HCW and SCD of 46% (95% CI: 31; 58) and 53% (95% CI: 32; 68), respectively. In 65PHC, significant VE’s were observed for all time periods after vaccination, but with increasing VE after the first dose (Table 2). In all priority groups, the incidence rates were highest in the unvaccinated group, except in LTCF residents and HCW, who had higher incidence rates the first 0-14 days after first dose, which resulted in negative VE estimates of -37% (95% CI: -55; -21) and -113% (95% CI: - 127; -101), respectively (Table 2). The overall stratified VE’s for the five priority groups were significant for the time periods 0-7 days after the second dose and >7 days after the second dose reaching 42% (95% CI: 33; 50) and 82% (95% CI: 79; 84), respectively.

### VE against all-cause hospital admission

For LTCF residents, 65PHC, +85, and HCW, significantly positive adjusted VE estimates were observed in all time-periods (Table 3). In addition, these VE estimates were similar across the four priority groups and ranged from 27% (95% CI: 18; 35) to 50% (95% CI: 45; 55). In the priority group SCD, significant positive VE estimates were observed until 7 days after the second dose, but, these VE estimates were generally lower compared to the other four priority groups ranging from 13% (95% CI: 7; 18) to 18% (95% CI: 14; 22). In the time-period >7 days after the second dose, significant negative adjusted VE estimates were observed of - 10% (95% CI: -17; -3) (Table 3). The overall VE’s against all-cause hospital admission for all five priority groups ranged from 21% (95% CI: 18; 23) to 30% (95% CI: 27; 33) (Table 3)

**Table 3.**
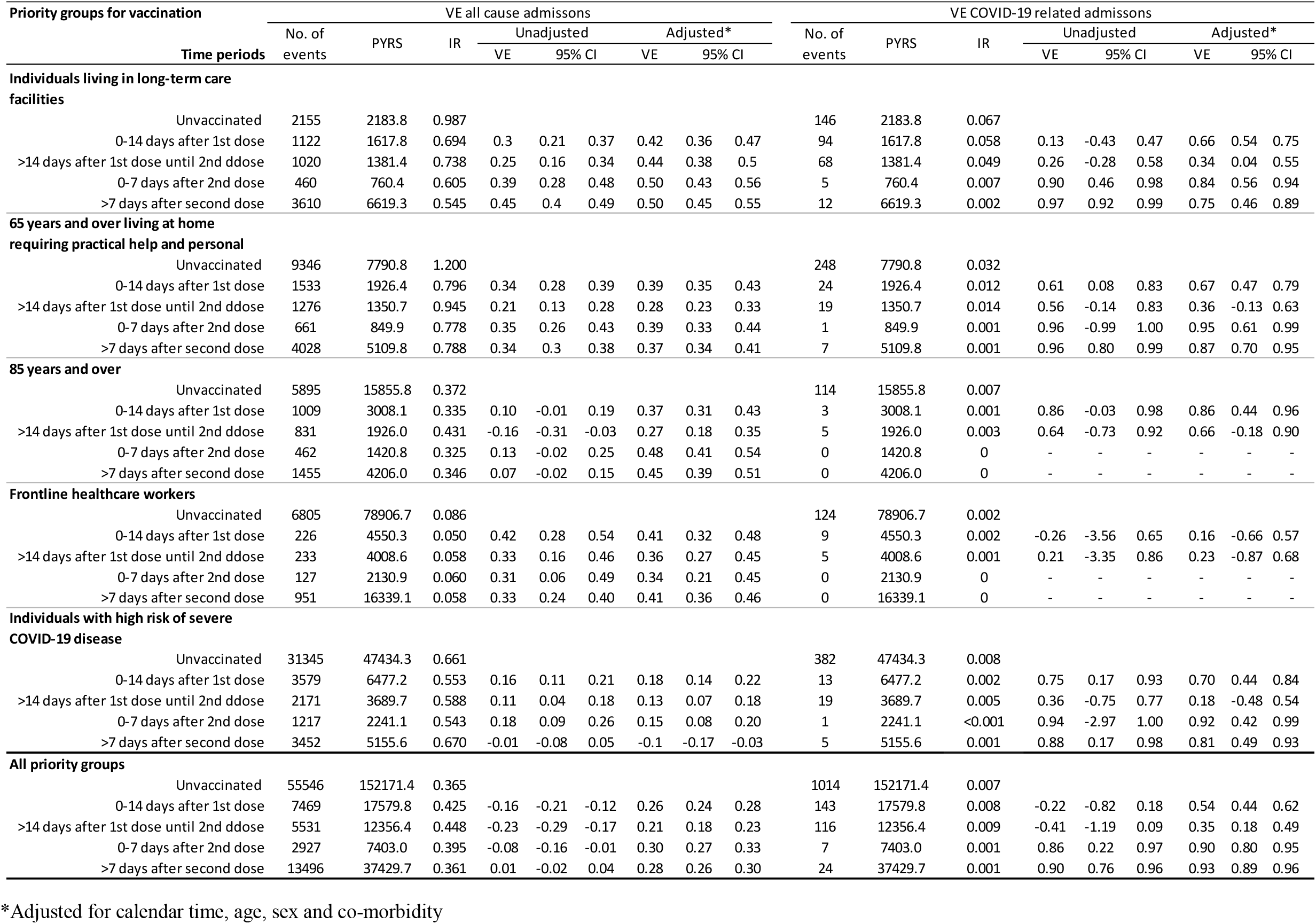
BNT162b2 mRNA vaccine effectiveness on all-cause hospital admissions and admissions related to COVID-19 in the priority groups long term care facility residents, individuals 65 years and above requiring practical help and personal care, 85 years and over, frontline healthcare workers, individuals with high risk of severe COVID-19 disease and in all priority groups combined.

### VE against hospital admission related to COVID-19

In LTCF residents, 65PHC, and SCD, a decrease in incidence rates was observed from unvaccinated to >7 days after the second dose. In all time periods, the highest incidence rates were observed in LTCF residents. In this group, significant VE estimates were seen in all time periods with the highest VE estimates 0-7 days after the second dose 84% (95% CI: 56; 94) and >7 days after the second dose 75% (95% CI: 46; 89). In 65PHC and SCD, significant VE’s were observed 0-14 days after the first dose, 0-7 days and >7 days after the second dose. More than 7 days after the second dose, the estimated VE’s were 87% (95% CI: 70; 95) in the 65PHC and 81% (95% CI: 49; 93) in the SCD priority groups (Table 3). There were very few admissions related to COVID-19 in the priority groups +85 and HCW and none after the second dose. Thus, VE could either not be estimated or was not significant except for +85 during the first 14 days after the first dose where VE was 86% (95% CI: 44; 96) (Table 3). The overall VE’s for the five priority groups were significant for all time periods and reached 90% (95% CI: 80; 95) 0-7 days after the second dose and 93% (95% CI: 89; 96) >7 days after the second dose.

### VE against all-cause death

In LTCF residents, 65PHC, +85, and SCD, the adjusted all-cause death VE’s were significantly positive in all time periods except from more than 14 days after the first dose until the second dose. Here, a negative VE estimate was observed for LTCF residents of -23% (95% CI: -39; -9) and a non-significant VE estimate for SCD of 13% (95% CI: -1; 25) (Table 4). In HCW, no significant vaccine effect in preventing all-cause death was observed (Table 4). The overall stratified VE’s against all-cause death for all five priority groups ranged from 6% (95% CI: 0; 12) to 66% (95% CI: 61; 70) (Table 4)

**Table 4.** BNT162b2 mRNA vaccine effectiveness on death as all cause death and COVID-19 related death in the priority groups long term care facility residents, individuals 65 years and above requiring practical help and personal care, 85 years and over, frontline healthcare workers, individuals with high risk of severe COVID-19 disease and in all priority groups combined.

| Priority groups for vaccination | Time period | VE all cause death |  |  |  |  |  |  |  |  | VE COVID-19 related death |  |  |  |  |  |  |  |  |
| --- | --- | --- | --- | --- | --- | --- | --- | --- | --- | --- | --- | --- | --- | --- | --- | --- | --- | --- | --- |
|  |  | No. of events | PYRS | IR | Unadjusted |  |  | Adjusted* |  |  | No. of events | PYRS | IR | Unadjusted |  |  | Adjusted* |  |  |
|  |  |  |  |  | VE | 95% CI |  | VE | 95% CI |  |  |  |  | VE | 95% CI |  | VE | 95% CI |  |
| Individuals living in long-term care facilities |  |  |  |  |  |  |  |  |  |  |  |  |  |  |  |  |  |  |  |
|  | Unvaccinated | 939 | 2232.2 | 0.421 |  |  |  |  |  |  | 118 | 2232.2 | 0.053 |  |  |  |  |  |  |
|  | 0-14 days after 1st dose | 446 | 1629.2 | 0.274 | 0.35 | 0.22 | 0.46 | 0.38 | 0.29 | 0.45 | 62 | 1629.2 | 0.038 | 0.28 | -0.26 | 0.59 | 0.77 | 0.68 | 0.83 |
|  | >14 days after 1st dose until 2nd ddose | 837 | 1400.5 | 0.598 | -0.42 | -0.66 | -0.22 | -0.23 | -0.39 | -0.09 | 162 | 1400.5 | 0.116 | -1.19 | -2.37 | -0.42 | -0.02 | -0.35 | 0.23 |
|  | 0-7 days after 2nd dose | 130 | 764.1 | 0.170 | 0.60 | 0.45 | 0.70 | 0.63 | 0.54 | 0.70 | 0 | 764.1 | 0 | - | - | - | - | - | - |
|  | >7 days after second dose | 1989 | 6668.1 | 0.298 | 0.29 | 0.19 | 0.38 | 0.26 | 0.17 | 0.34 | 23 | 6668.1 | 0.003 | 0.93 | 0.85 | 0.97 | 0.89 | 0.81 | 0.93 |
| >65 years living at home requiring practical help and personal care |  |  |  |  |  |  |  |  |  |  |  |  |  |  |  |  |  |  |  |
|  | Unvaccinated | 2134 | 7988.5 | 0.267 |  |  |  |  |  |  | 170 | 7988.5 | 0.021 |  |  |  |  |  |  |
|  | 0-14 days after 1st dose | 192 | 1943.5 | 0.099 | 0.63 | 0.52 | 0.72 | 0.59 | 0.52 | 0.65 | 4 | 1943.5 | 0.002 | 0.9 | 0.38 | 0.98 | 0.94 | 0.84 | 0.98 |
|  | >14 days after 1st dose until 2nd ddose | 360 | 1379.8 | 0.261 | 0.02 | -0.19 | 0.2 | 0.21 | 0.11 | 0.30 | 26 | 1379.8 | 0.019 | 0.11 | -0.92 | 0.59 | 0.02 | -0.54 | 0.37 |
|  | 0-7 days after 2nd dose | 60 | 855.5 | 0.070 | 0.74 | 0.58 | 0.83 | 0.70 | 0.61 | 0.77 | 0 | 855.5 | 0 | - | - | - | - | - | - |
|  | >7 days after second dose | 600 | 5173.3 | 0.116 | 0.57 | 0.49 | 0.63 | 0.62 | 0.57 | 0.66 | 2 | 5173.3 | 0.0004 | 0.98 | 0.75 | 1.00 | 0.97 | 0.88 | 0.99 |
| 85 years and over |  |  |  |  |  |  |  |  |  |  |  |  |  |  |  |  |  |  |  |
|  | Unvaccinated | 1333 | 15947.4 | 0.084 |  |  |  |  |  |  | 75 | 15947.4 | 0.005 |  |  |  |  |  |  |
|  | 0-14 days after 1st dose | 78 | 3017.6 | 0.026 | 0.69 | 0.52 | 0.8 | 0.67 | 0.58 | 0.75 | 0 | 3017.6 | 0 | - | - | - | - | - | - |
|  | >14 days after 1st dose until 2nd ddose | 130 | 1941.3 | 0.067 | 0.20 | -0.14 | 0.44 | 0.50 | 0.38 | 0.60 | 8 | 1941.3 | 0.004 | 0.12 | -1.89 | 0.73 | -0.15 | -8.22 | 0.86 |
|  | 0-7 days after 2nd dose | 27 | 1424.0 | 0.019 | 0.77 | 0.52 | 0.89 | 0.80 | 0.70 | 0.87 | 0 | 1424.0 | 0 | - | - | - | - | - | - |
|  | >7 days after second dose | 142 | 4225.4 | 0.034 | 0.60 | 0.44 | 0.71 | 0.73 | 0.66 | 0.79 | 0 | 4225.4 | 0 | - | - | - | - | - | - |
| Frontline healthcare workers |  |  |  |  |  |  |  |  |  |  |  |  |  |  |  |  |  |  |  |
|  | Unvaccinated | 73 | 78971.5 | 0.001 |  |  |  |  |  |  | 3 | 78971.5 | 0.0004 |  |  |  |  |  |  |
|  | 0-14 days after 1st dose | 4 | 4552.3 | 0.001 | 0.05 | -11.32 | 0.93 | 0.46 | -0.51 | 0.80 | 0 | 4552.3 | 0 | - | - | - | - | - | - |
|  | >14 days after 1st dose until 2nd ddose | 3 | 4011.3 | 0.001 | 0.19 | -14.28 | 0.96 | 0.47 | -0.71 | 0.84 | 0 | 4011.3 | 0 | - | - | - | - | - | - |
|  | 0-7 days after 2nd dose | 1 | 2131.8 | 0.001 | 0.49 | -76.03 | 1.00 | 0.69 | -1.28 | 0.96 | 0 | 2131.8 | 0 | - | - | - | - | - | - |
|  | >7 days after second dose | 12 | 16348.4 | 0.001 | 0.21 | -2.76 | 0.83 | 0.23 | -0.54 | 0.62 | 0 | 16348.4 | 0 | - | - | - | - | - | - |
| Individuals with high risk of severe COVID-19 disease |  |  |  |  |  |  |  |  |  |  |  |  |  |  |  |  |  |  |  |
|  | Unvaccinated | 1940 | 48040.1 | 0.040 |  |  |  |  |  |  | 79 | 48040.1 | 0.002 |  |  |  |  |  |  |
|  | 0-14 days after 1st dose | 123 | 6524.5 | 0.019 | 0.53 | 0.31 | 0.69 | 0.62 | 0.54 | 0.68 | 3 | 6524.5 | 0.0005 | 0.72 | -2.98 | 0.98 | 0.77 | -1.2 | 0.98 |
|  | >14 days after 1st dose until 2nd ddose | 250 | 3738.0 | 0.067 | -0.66 | -1.2 | -0.25 | 0.13 | -0.01 | 0.25 | 7 | 3738.0 | 0.002 | -0.14 | -5.76 | 0.81 | 0.41 | -4.37 | 0.93 |
|  | 0-7 days after 2nd dose | 34 | 2254.4 | 0.015 | 0.63 | 0.22 | 0.82 | 0.72 | 0.60 | 0.80 | 0 | 2254.4 | 0 | - | - | - | - | - | - |
|  | >7 days after second dose | 209 | 5216.5 | 0.040 | 0.01 | -0.35 | 0.27 | 0.46 | 0.37 | 0.54 | 0 | 5216.5 | 0 | - | - | - | - | - | - |
| All priority groups |  |  |  |  |  |  |  |  |  |  |  |  |  |  |  |  |  |  |  |
|  | Unvaccinated | 6419 | 153179.6 | 0.042 |  |  |  |  |  |  | 445 | 153179.6 | 0.003 |  |  |  |  |  |  |
|  | 0-14 days after 1st dose | 843 | 17667.0 | 0.048 | -0.14 | -0.32 | 0.02 | 0.47 | 0.43 | 0.51 | 69 | 17667.0 | 0.004 | -0.34 | -1.42 | 0.25 | 0.76 | 0.68 | 0.82 |
|  | >14 days after 1st dose until 2nd ddose | 1580 | 12470.8 | 0.127 | -2.02 | -2.39 | -1.7 | 0.06 | 0.00 | 0.12 | 203 | 12470.8 | 0.016 | -4.6 | -7.24 | -2.81 | 0.07 | -0.15 | 0.25 |
|  | 0-7 days after 2nd dose | 252 | 7429.7 | 0.034 | 0.19 | -0.05 | 0.38 | 0.66 | 0.61 | 0.70 | 0 | 7429.7 | 0 | - | - | - | - | - | - |
|  | >7 days after second dose | 2952 | 37631.7 | 0.078 | -0.87 | -1.05 | -0.71 | 0.49 | 0.46 | 0.52 | 25 | 37631.7 | 0.001 | 0.77 | 0.42 | 0.91 | 0.94 | 0.90 | 0.96 |

\*Adjusted for calendar time, age, sex, co-morbidities and hospital admission

### VE against death related to COVID-19

Death related to COVID-19 infection was a rare event among vaccinated individuals in +85, HCW and SCD, and thus, VE could either not be estimated or was non-significant. In LTCF residents and 65PHC, significant VE’s against COVID-19 related death were observed 0-14 days after the first dose and >7 days after the second dose with VE estimates ranging from 77% (95% CI: 68; 83) to 97% (95% CI: 88; 99) (Table 4). The overall VE’s for the five priority groups were 76% (95% CI: 68; 82) 0-14 days after the first dose and 94% (95% CI: 90; 96) >7 days after the second dose.

## Discussion

We present BNT162b2 mRNA VE estimates in the five priority groups, who were vaccinated during the first 3½ months of the Danish COVID-19 vaccination programme. It is encouraging that among the most vulnerable citizens we observed a high reduction in risk of infection >7 days after the second dose. This includes VE estimates of 53% (95% CI: 29; 69) in LTCF residents, 86% (95% CI: 78; 91) in 65PHC, 77% (95% CI: 50; 89) in +85, and 71% (95% CI: 58; 80), in the SCD group. The estimated VE for LTCF residents was a little lower compared to the first preliminary Danish results reported by Moustsen-Helms et al. of (64% (95% CI: 14;84) [2]. In 65PHC, HCW and SCD, we observed a substantial reduction in risk of infection 0-7 days after the second dose (Table 2).

For +85 and SCD, no COVID-19 related deaths were observed after the second dose and for the +85, this was also the case for COVID-19 related admissions. In LTCF residents, 65PHC and SCD, VE against COVID-19 related admissions were above 80% already after the second dose was administered. Similarly, VE against COVID-19 related death was above 75% after receiving the second dose for LTCF residents and 65PHC. These significant protective VE estimates suggest that the BNT162b2 mRNA vaccine reduces the risk of admissions to hospital and death related to COVID-19. Dagan et al. [12] found that VE against COVID-19 related hospitalisation and death from 14 through 20 days after first dose was 74% (95% CI: 56; 86) and 72% (95% CI: 19-100), respectively, and beyond 7 days after the second dose, VE against hospitalisation was 87% (95% CI: 55-100) which is similar to our findings.

In contrast to the other priority groups, HCW represent healthy individuals. In this group, we found a significant reduction in the risk of infection from >7 days after the second dose of 80% (95% CI: 77; 83). This is similar to findings from England and the US where VE in this group was 85% (95% CI: 74; 96) [13] and 90% (95% CI: 68; 97) [14], respectively. In the latter study, VE was estimated ≥14 days after the second dose. Our findings in this study were slightly lower compared to the preliminary Danish results reported by Moustsen-Helms et al [2], where VE >7 days after the second dose was 90% (95% CI: 82; 95). No COVID-19 related admissions were observed among HCW after the second vaccine dose and no COVID-19 related deaths were observed after the first vaccine dose hence VE for these outcomes could not be estimated.

LTCF residents are among the most vulnerable individuals in the population and combined with a high age distribution, this is the most likely reason why several countries have put this group first in line to receive the COVID-19 vaccine [2,15,16]. However, few studies have reported VE for this group. In a study on LTCF residents from Connecticut, Britton et al. [15] reported a BNT162b2 mRNA VE of 63% (95% CI: 33; 79) from more than 14 days after the first dose to 7 days after the second dose, and a VE of 60% (95% CI: 33; 77) 14 days after the second dose, which is comparable to our findings of 53% (95% CI: 29; 69) for this group. In Denmark, the first and second dose BNT162b2 mRNA vaccine coverage increased among the LTCF residents and within a couple of weeks, the majority had received the first dose and the same was seen with the second dose, thus giving a short time period with overlap between unvaccinated and vaccinated for both doses. Thereby, a major part of the comparison was based on few and maybe selected persons. This implies that the VE estimates in LTCF residents should be interpreted with some caution. A similar scenario was also reported by Britton et al. [15].

The significant negative effect of the vaccine observed in LTCF residents and HCW the first 0-14 days after the first dose was not unexpected. These priority groups received the vaccine when it became available at the end of December 2020 and at that time there were outbreaks at several LTCF and a high incidence of SARS-CoV-2 in the population. Therefore, we cannot rule out that some LTCF residents and HCW were infected at the time of vaccination, or shortly after they received the first dose. This increases the risk that many SARS-CoV-2 cases were counted as infected after receiving the first dose even though the infection happened prior to vaccination or before immunity. Similar observations were reported in a UK study [17].

In a UK study, Bernal et al. reported a VE of 89% (95% CI 85; 93) among individuals ≥80 years of age 14 days after the second dose [16], which is higher than our finding for the +85, where we estimated a VE of 77% (95% CI: 50; 89). Similar to LTCF residents, a steep increase in the first and second of BNT162b2 mRNA vaccine coverage was observed in the priority group +85 (Figure 1), and therefore a major part of the comparison might have been based on few and maybe selected persons. This implies that similarly to the VE estimates from the LTCF residents, the VE estimates from the +85 should be interpreted with some caution.

In Denmark, a large effort was done to ensure that all individuals in the priority groups were offered the COVID-19 vaccines and had the same probability of receiving them. However, the significant protective VE estimates on all-cause admissions and all-cause death in almost all priority groups and time periods indicate that the vaccinations were postponed in ill individuals. Postponed vaccinations due to illness e.g. COVID-19, could increase the number of events under investigation among unvaccinated. This could result in an underrepresentation of ill individuals during the first days after vaccination, which will imply that the VE in a period after vaccination may be artificially increased. This might explain the high VE estimates 0-14 days after vaccination compared with the time period from >14 days after vaccination until the second dose, which is observed for COVID-19 related admissions in LTCF residents, 65PHC, +85, and SCD and for COVID-19 related death among LTCF residents, 65PHC, and SCD.

The significant negative VE on all-cause admission to hospital >7 days after the second dose in the SCD group was unsurprising as these individuals were identified due to their underlying diseases, and the COVID-19 vaccine was not expected to prevent hospital admissions caused by diseases not related to COVID-19.

In the priority groups 65PHC, HCW, and SCD, we observed an effect of the vaccine from 0-7 days after the second dose. In several studies, this time period is included as an effect of the first dose. We have analysed this time period separately to insure that the effect of the first and second dose were not mixed. A similar approach was used by Thompson et al [14].

### Strengths

This study has several strengths. Our five priority groups are national cohorts and all the information that we link, including COVID-19 vaccinations, laboratory confirmed SARS-CoV-2 infections, co-morbidities, vital status, age, sex and hospital admission, are national data available at individual level and in real time. In Denmark, healthcare, including SARS-CoV-2 test and vaccination, is free of charge, and Denmark has a national strategy, where all individuals can be tested for SARS-CoV-2 and have an answer within 24 hours. Rapid antigen tests and RT-PCR tests are available and recorded in MiBa, however a positive antigen test must be verified by a PCR test. During the study period, HCWs in hospital were offered weekly testing.

### Limitations

End of December 2020 beginning of January 2021, the SARS-CoV-2 incidence in the population was very high and COVID-19 outbreaks were observed at several LTCF [2]. As a result, most of the COVID-19 events were observed during the start of the study, where a large proportion of the priority groups were unvaccinated. Therefore, it was important to account for calendar time in the estimation of the VE’s. In all priority groups, the second dose was administered at a median of 22-25 days after first dose, which did not allow sufficient time to observe the full effect of the first dose. With a high vaccine coverage as observed among LTCF and 85+, the unvaccinated group that is used for comparison becomes very small and include a selected population different from the majority. This implies that VE estimates in this setting should be interpreted with caution.

In conclusion, across all five priority groups, the overall VE against COVID-19 infection >7 days after the second dose was 82%. It is encouraging that among the most vulnerable citizens and those with the highest risk of infection, we observed a high reduction in risk of infection when fully vaccinated with BNT162b2 mRNA. In addition, though COVID-19 related death in many priority groups are rare events following vaccination, over all COVID-19 related admissions and deaths were reduced by 93% and 94%, respectively.

## Data Availability

The individual level data used in this study are sensitive and cannot be publicly shared. Any data requests should be sent to Forskerservice at the Danish Health and Medicin Authority. Aggregated surveillance data can are freely available online: https://covid19.ssi.dk/

## Ethical considerations

The present study was based on existing administrative data and did not require ethical approval

## Role of funding source

Neither Statens Serum Institut nor any authors received funding for this study.

## Declaration of interest

All authors declared no competing interest

## Acknowledgement

The authors are grateful to the Danish Health Data Authority for their help defining the population cohorts. We would also like to thank the Department of Data Integration and Analysis at Statens Serum Institut for the data management.

## Notes

### Competing Interest Statement

The authors have declared no competing interest.

### Author Declarations

The data included in this report is part of the Danish national COVID-19 surveillance system database at Statens Serum Institut. The data becomes or are already available for research upon reasonable request and with permission from the Danish Data Protection Agency and Danish Health and Medicines Authority. Ethics: We used only administrative register data for the study. According to Danish law, ethics approval is exempt for such research, and the Danish Data Protection Agency, which is a dedicated ethics and legal oversight body, thus waives ethical approval for our study of administrative register data, when no individual contact of participants is neccessary and only aggregate results are included as findings. The study is therefore fully compliant with all legal and ethical requirements and there are no further processes available regarding such studies.

